# Retinal Vascular Parameters and Risk of Heart Failure

**DOI:** 10.1101/2023.05.31.23290813

**Authors:** Yuting Wu, Yueye Wang, Qi Chen, Ruobing Wang, Mingguang He, Danli Shi

## Abstract

**Aim:** To investigate the association of retinal vascular parameters (calibre, tortuosity, fractal dimension and the number of segments) with incident heart failure (HF) in the UK Biobank.

**Methods:** Participants with fundus images in UK Biobank were included. Retinal vascular parameters were assessed by an artificial intelligence system (RMHAS). Incident HF events were determined by hospital inpatient data, death register records and self-reported medical condition. Cox proportional hazards regression models (CPH) were used to investigate the links between retinal vascular variables and the risk of HF.

**Results:** We included 56470 individuals. After a median of 10.95-year follow-up, 1062 incident HF events were reported. After adjusting for conventional HF risk factors in CPH, we found larger retinal venular calibre (hazard ratio [HR], 1.54; 95% confidence interval [CI], 1.12-2.14), smaller arteriolar fractal dimension (HR, 1.98; 95% CI, 1.38-2.85), smaller venular fractal dimension (HR, 1.52; 95% CI, 1.09-2.13), the smaller number of arteriolar segments (HR, 2.18; 95% CI, 1.47-3.24) and the smaller number of venular segments (HR, 1.85; 95% CI, 1.29-2.64) were linked to an increased risk of HF. The addition of retinal vessel measurements to traditional HF risk factors increased the discrimination capability for predicting future HF, with C statistics changing from 0.799 to 0.813.

**Conclusions:** Wider retinal venular calibre, lower vascular fractal dimension and a smaller number of vessel segments were associated with a higher risk of incident HF events. These results point to the potential application of using early retinal vasculature changes to identify people at higher risk of HF.

## Introduction

Heart failure (HF), the most rapidly growing cardiovascular condition globally^1^, is a progressive and life-threatening clinical condition affecting more than 60 million people worldwide, conferring a substantial burden on healthcare systems worldwide^2,3^. The early symptoms of HF are not obvious, but the condition becomes irreversible at the late stage despite the availability of treatment^4^. Early detection and intervention can effectively reduce the incidence of HF and mortality.

It has been demonstrated that microvascular disorders predominately contribute to the development of HF^5^. Microcirculation disturbance directly affects microcirculation perfusion, further causing the corresponding coronary artery disease, thus aggravating the deterioration of heart function, eventually leading to HF^6^. The retina offers a unique opportunity to directly evaluate microvasculature in a noninvasive way, which may provide a unique view of systemic microcirculation. Mounting evidence has identified the close association between retinal vascular changes and the risk of developing various cardiovascular diseases ^7,8^.

With the development of retinal imaging and vascular segmentation algorithms, increasing studies were able to more efficiently evaluate the association between quantified retinal vasculature and cardiovascular events. Several studies have found that different types of heart failure were associated with retinopathy and retinal vascular parameters (e.g. retinal arteriolar calibre, central retinal arteriolar equivalents (CRAE), central retinal venular equivalents (CRVE))^9-12^. It has also been demonstrated that some morphological retinal vascular characteristics, such as tortuosity and vasculature fractal, which represent the branching network and complexity of the retinal vasculature, are connected to stroke and cardiovascular disorders^13-15^. However, current studies only measured limited exposures, mainly on calibre-related parameters, with few performing a comprehensive assessment of retinal vascular characteristics. Furthermore, there is limited evidence on whether digital retinal microvascular assessment will provide any further advantages for HF risk differentiation over the conventional risk factors remains to be seen. Recently, in order to gain a more thorough understanding of the retinal vessels, we have developed an artificial intelligence system (Retina-based Microvascular Health Assessment System, RMHAS)^16^ for automated vessel segmentation and quantification of a wide range of retinal vessel parameters.

Taken together, this study aimed to investigate the relationships between a comprehensive set of retinal vascular characteristics (calibre, tortuosity, fractal dimension and the number of segments) and the risk of developing heart failure in the UK Biobank population. We also examined the cumulative impact of standard risk variables for HF prediction and retinal microvascular parameters.

## Methods

### Study Population

Data for this analysis was derived from the UK Biobank study, a large-scale and prospective cohort study of 502,656 UK population aged between 40 to 70 years who were registered with the National Health Service (NHS)^17^. Between January 2006 and October 2010, baseline questionnaires and physical measurements were taken at 22 evaluation centres. An initial ophthalmic examination was performed between June 2009 and July 2010. Later, over 20 000 participants underwent follow-up ophthalmic tests from August 2012 to June 2013^18^. Retinal fundus images were taken from a subset of 83,573 participants, yielding 172,899 retinal photographs (from both eyes). The overall protocol is available online (http://www.ukbiobank.ac.uk/resources/). The UK Biobank received ethics approval from the North West Research Ethics Committee (reference number 06/MRE08/65). All participants provided informed consent. The study was performed following the principles of the Declaration of Helsinki.

### Assessment of Retinal Microvasculature

The retina was photographed in colour images using a Topcon 3D OCT-1000 Mark II device. The detailed methodology has been described previously^19^.

We employed a deep learning system, the RMHAS^16^, to quantitatively evaluate the retinal microvascular characteristics from digital retinal photographs using completely automated vessel segmentation (Figure 1). Measurements were taken of all visible vessels in the designated area. Using the RMHAS program, a number of retinal vascular variables were measured and a panel of retinal vascular measurements was generated automatically, including calibre, tortuosity, fractal dimension, and the number of vascular segments. Arteriole and venular were measured separately. Retinal calibre was the mean of diameters of all identified vessel segments. The tortuosity of retinal vessels was calculated by segment length along the centre line divided by linear endpoint distance. Fractal dimension was calculated by the box-counting method, expressing the density and overall complexity of retinal vasculature in one number.

**Figure 1.**
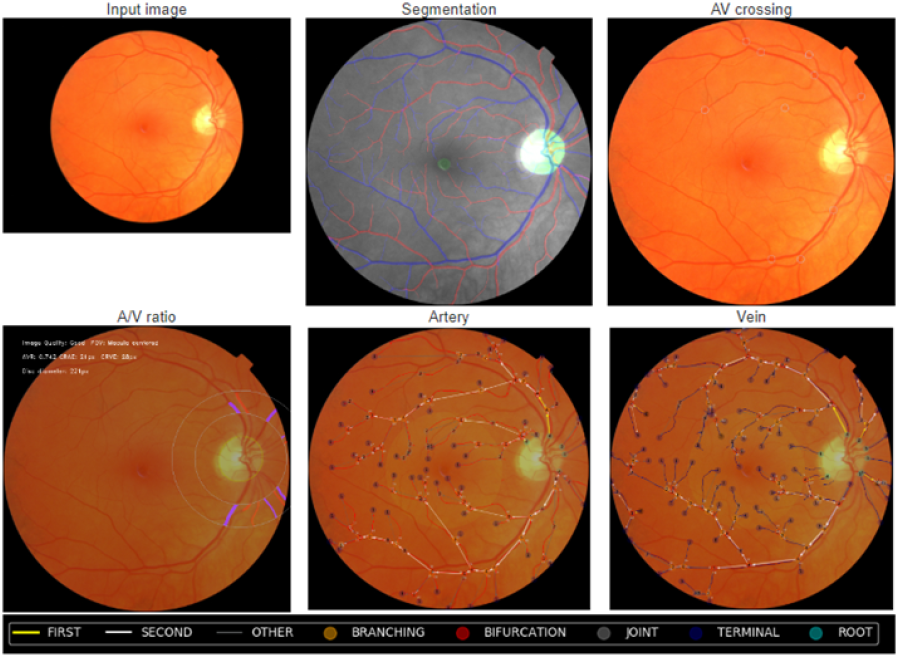
Retinal fundus photographs assessed quantitatively by RMHAS software.

### Assessment of Heart Failure Outcome

Incident heart failure from all causes that first occurred during the period between baseline examination and April 28, 2021, was recorded in the UK Biobank study by hospital data codes (International Classification of Diseases, 9th Revision, Clinical Modification (ICD-9-CM); International Classification of Diseases, 10th Revision (ICD-10)) and self-reported data codes (UKB data coding 6). A hospitalization case would be considered to have a heart failure event if it contains a hospital discharge diagnosis code of heart failure (ICD-9-CM code 428 or 518.4 or ICD-10 code I50). Self-reported data code of heart failure is 1076. The date of heart failure’s first occurrence was generated by mapping hospital inpatient data, death register records, and self-reported medical condition codes collected at the baseline or subsequent UK Biobank assessment centre visit, whichever came first.

### Assessment of Risk Factors

Participants underwent baseline questionnaires and physical measurements. A computerized questionnaire was used to collect data on age, sex, ethnicity, smoking status, and medicine usage. Weight (in kilograms) divided by squared height (in meters) was used to calculate BMI. Blood pressure was defined as the mean of two repeated measures with at least a 1-minute interval taken with a HEM-7015IT electronic blood pressure monitor (Omron Healthcare, the Netherlands) in a seated position. Five Variant II Turbo analyzers (Bio-Rad Laboratories, USA) were applied to conduct the glycated hemoglobin (HbA1c) assay. Plasma glucose, triacylglycerol (TG), C-reactive protein (CRP), total cholesterol (TC), high-density lipoprotein cholesterol (HDL-c), and C-reactive protein (CRP) were also tested. Ten immunoassay analysers (six Liaison XL DiaSorin Italy and four DXI 800 analysers, Beckman Coulter Diagnostics, Switzerland) and four clinical chemistry analysers (two AU5800 Beckman Coulter Diagnostics and two Advia 1800 analysers Siemens, Germany) were utilized for the analyses of serum biomarkers^20^. At the baseline assessment, a history of CHD, stroke, diabetes, and hypertension were identified by hospital inpatient data and self-reporting questionnaires.

### Statistical Analysis

Sex-specific extreme outliers were excluded by adjusting the traditional box and whisker upper and lower bounds and accounting for skewness using the Robustbase package in R (setting range=3)^7^.

Independent t-tests, Mann-Whitney U tests, or chi-square tests were used to examine the differences between participants with and without incident HF. Baseline characteristics were summarised using descriptive statistics for continuous variables (mean ± standard deviation or medium (IQR)) and numbers (percentages) for categorical variables. All retinal microvascular parameters were analysed continuously and then classified into four quartiles for categorical assessment.

Cox proportional-hazards regression models were used to investigate the links between RMHAS-measured retinal vascular variables and the risk of HF. Covariates were chosen according to traditional risk factors described previously in the literature^21^. All Cox regression analyses were based on four models. Model 1 includes the age and sex. Model 2 adjusted for traditional risk factors (age, sex, systolic blood pressure, total cholesterol, high-density lipoprotein cholesterol, triglyceride, body mass index, smoking, and use of antihypertensive medication). Model 3 includes established risk factors and was further adjusted for diabetic retinopathy and retinal vessel occlusion (RVO). In addition to traditional risk factors, Model 4 additionally adjusted for a history of stroke and a history of CHD. HRs for continuous variables relate to a 1 SD increase or 1SD decrease. Prespecified subgroup analyses were performed after stratification by gender, hypertension, diabetes, history of stroke, and history of CHD.

To evaluate the model’s predictive accuracy, we used concordance statistics (or C statistics) to assess the discriminative capability of the risk-factor-based model and the model combining traditional HF risk factors and retinal vessel parameters. R 3.5.1 was used to conduct all of the analyses. Two-sided P-values of 0.05 were deemed significant if not otherwise indicated.

## Result

A total of 56470 participants were included in the analysis, and the inclusion and exclusion flow chart is shown in Figure 2. We excluded the subjects whose fundus images failed quality control, participants who lost follow-up or had a history of HF, and individuals with missing data. The median follow-up time was 10.95 years (IQR 10.82-11.10). Table 1 provides a summary of the baseline demographic profile of the included participants. Overall, 25480 participants (45.1%) were male, with an average age of 56.61 at baseline. We also compared the baseline variables with participants whether or not had an incident heart failure. All covariates except for diastolic blood pressure were found to have significant differences between the two groups of participants whether or not had an incident HF. Individuals with a history of diabetic retinopathy, retinal vessel occlusion, stroke, and CHD were more likely to have prevalent HF. Details are described in Table 1.

**Table 1.** Baseline characteristics of participants with incident heart failure and no events

| Variable | All<br>(N=56470) | Incident HF<br>(N=1062) | No incident HF<br>(N=55408) | *P* value |
| --- | --- | --- | --- | --- |
| Age (years) | 56.73±8.23 | 62.84±6.07 | 56.61±8.22 | <0.001 |
| Male, n (%) | 25480 (45.1) | 661 (62.2) | 24819 (44.8) | <0.001 |
| Ethnicity |  |  |  | 0.177 |
| White | 51701 (91.6) | 978 (92.1) | 50723 (91.5) |  |
| Other | 4769 (8.4) | 84 (7.9) | 4685 (8.5) |  |
| SBP, mmHg | 136.78±18.26 | 142.85±19.16 | 136.66±18.22 | <0.001 |
| DBP, mmHg | 81.49±10.00 | 81.80±10.51 | 81.48±9.99 | 0.306 |
| BMI, kg/m<sup>2</sup> | 27.20±4.72 | 29.51±5.57 | 27.16±4.69 | <0.001 |
| Total cholesterol, mmol/L | 5.70±1.08 | 5.22±1.15 | 5.71±1.08 | <0.001 |
| HbA1c, mmol/mol | 35.98±6.04 | 39.18±10.59 | 35.91±5.90 | <0.001 |
| HDL-c, mmol/L | 1.49±0.36 | 1.38±0.35 | 1.49±0.36 | <0.001 |
| LDL-c, mmol/L | 3.54±0.82 | 3.21±0.86 | 3.55±0.82 | <0.001 |
| Triglyceride, mmol/L | 1.67±0.91 | 1.66±0.91 | 1.79±1.01 | <0.001 |
| BP medications, n (%) | 10901 (19.3) | 526 (49.5) | 10375 (18.7) | <0.001 |
| Hypertension | 29181 (51.7) | 835 (78.6) | 28346 (51.2) | <0.001 |
| Diabetes mellitus | 3135 (5.6) | 179 (16.9) | 2956 (5.3) | <0.001 |
| Current smoking, n (%) | 4899 (8.7) | 139 (13.1) | 4760 (8.6) | <0.001 |
| Diabetic retinopathy, n (%) | 408 (0.7) | 59 (5.6) | 349 (0.6) | <0.001 |
| RVO, n (%) | 175 (0.3) | 15 (1.4) | 160 (0.3) | <0.001 |
| History of stroke | 824 (1.5) | 50 (4.7) | 774 (1.4) | <0.001 |
| History of CHD | 2586 (4.6) | 263 (24.8) | 2323 (4.2) | <0.001 |

Summary statistics for independent variables were calculated as means and standard deviation (SD) or median and interquartile range (IQR) for continuous variables, case numbers, and percentages for categorical variables.
SBP: systolic blood pressure; DBP: diastolic blood pressure; BMI: body mass index; HbA1c: glycated haemoglobin; HDL-c: high-density lipoprotein cholesterol; LDL-c: low-density lipoprotein cholesterol; BP medications: blood pressure medications; RVO: retinal vascular occlusion; CHD: coronary heart disease

**Figure 2.**
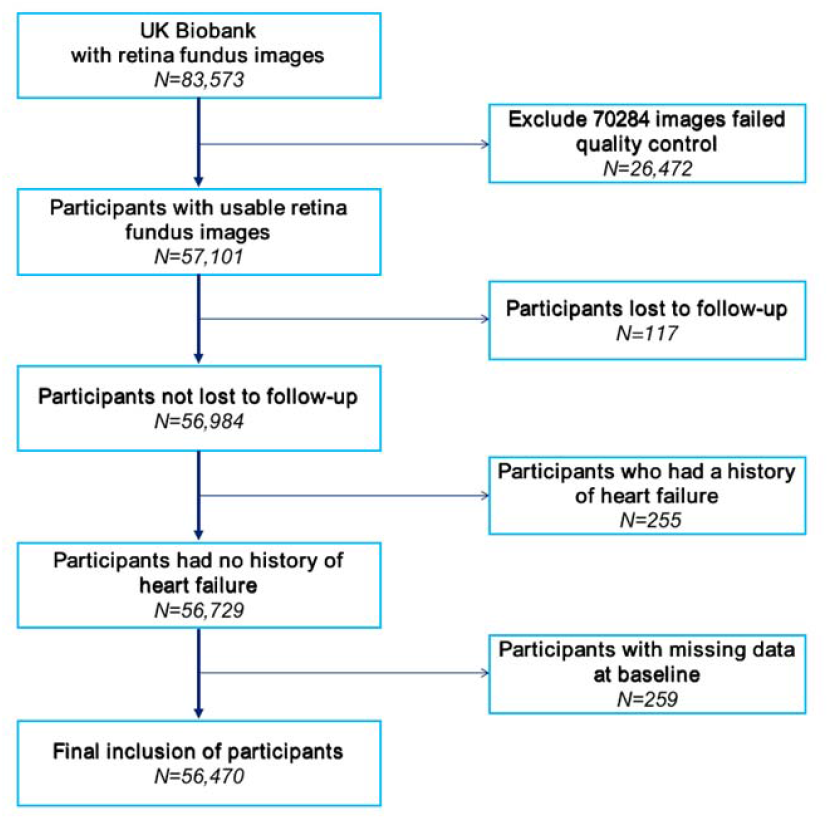
Inclusion and exclusion flow chart.

### Association between retinal vascular parameters and incident HF

The HRs of specific retinal vascular parameters linked to the risk of HF were listed in Table 2. In Cox proportional-hazards models adjusting for traditional risk factors (age, sex, systolic blood pressure, TC, HDL-c, TG, BMI, smoking and use of antihypertensive medication), wider retinal venous calibre (HR, 1.28; 95% CI, 1.06-1.54; *P*=0.009) categorized in the fourth quartile was associated with a higher risk of HF, compared to the vessel calibre in the first quartile. When modeled as a continuous variable, increased retinal venous calibre was associated with a higher HF incidence (HR,1.54; 95% CI, 1.12-2.14; *P*=0.009; per SD increase). After further adjustment for traditional risk factors and retinopathy (diabetic retinopathy and RVO), those in the fourth quartile of the retinal venous calibre, relative to the first quartile, presented a 27% higher risk of HF (HR, 1.27; 95% CI, 1.06-1.53; *P*=0.011). Such associations remained significant, along with further adjustment for a history of stroke and CHD. Interestingly, for arterial calibre, significant results were found as a categorical variable (Model 2: HR, 0.81; 95% CI, 0.67-0.99; *P*=0.036. Model 3: HR, 0.81; 95% CI, 0.66-0.98; *P*=0.033. Model 4: HR, 0.81; 95% CI, 0.67-0.99; *P*=0.036, comparing second versus first quartiles). Compared with the first quartile, the risk of HF was reduced by 19% in the second quartile. If only adjusting for age and sex, the increased retinal arteriolar diameter was associated with increased HF incidence (HR, 1.57; 95% CI, 1.11-2.22; *P*=0.011; per SD increase).

**Table 2.** Relationship of selected major retinal vascular parameters with risk of heart failure

| Parameters | Model 1 |  | Model 2 |  | Model 3 |  | Model 4 |  |
| --- | --- | --- | --- | --- | --- | --- | --- | --- |
|  | HR (95% CI) | P Value | HR (95% CI) | P Value | HR (95% CI) | P Value | HR (95% CI) | P Value |
| Retinal arteriolar calibre |  |  |  |  |  |  |  |  |
| Q1 (n=13541) | Reference | - | Reference | - | Reference | - | Reference | - |
| Q2 (n=13538) | 0.84 (0.69, 1.03) | 0.089 | 0.81 (0.67, 0.99) | 0.036 | 0.81 (0.66, 0.98) | 0.033 | 0.81 (0.67, 0.99) | 0.036 |
| Q3 (n=13539) | 1.13 (0.94, 1.35) | 0.197 | 1.05 (0.87, 1.25) | 0.622 | 1.05 (0.87, 1.25) | 0.631 | 1.03 (0.86, 1.24) | 0.722 |
| Q4 (n=13540) | 1.07 (0.89, 1.28) | 0.489 | 0.98 (0.82, 1.18) | 0.854 | 0.96 (0.80, 1.15) | 0.654 | 0.97 (0.81, 1.16) | 0.703 |
| Per SD increase | 1.57 (1.11, 2.22) | 0.011 | 1.34 (0.94, 1.91) | 0.103 | 1.28 (0.90, 1.82) | 0.169 | 1.28 (0.90, 1.83) | 0.165 |
| Retinal venular calibre |  |  |  |  |  |  |  |  |
| Q1 (n=13540) | Reference | - | Reference | - | Reference | - | Reference | - |
| Q2 (n=13539) | 1.05 (0.86, 1.29) | 0.613 | 1.02 (0.83, 1.25) | 0.859 | 1.02 (0.84, 1.25) | 0.829 | 1.01 (0.83, 1.24) | 0.913 |
| Q3 (n=13539) | 1.25 (1.03, 1.51) | 0.021 | 1.14 (0.94, 1.38) | 0.178 | 1.14 (0.94, 1.38) | 0.169 | 1.13 (0.93, 1.37) | 0.214 |
| Q4 (n=13540) | 1.45 (1.21, 1.74) | <0.001 | 1.28 (1.06, 1.54) | 0.009 | 1.27 (1.06, 1.53) | 0.010 | 1.27 (1.06, 1.53) | 0.011 |
| Per SD increase | 1.95 (1.41, 2.68) | <0.001 | 1.54 (1.12, 2.14) | 0.009 | 1.51 (1.09, 2.08) | 0.014 | 1.52 (1.09, 2.10) | 0.012 |
| Retinal arteriolar tortuosity |  |  |  |  |  |  |  |  |
| Q1 (n=13386) | Reference | - | Reference | - | Reference | - | Reference | - |
| Q2 (n=13375) | 0.94 (0.79, 1.12) | 0.489 | 0.95 (0.79, 1.12) | 0.525 | 0.93 (0.78, 1.11) | 0.428 | 0.94 (0.79, 1.12) | 0.507 |
| Q3 (n=13378) | 0.92 (0.77, 1.09) | 0.322 | 0.91 (0.77, 1.09) | 0.307 | 0.91 (0.76, 1.08) | 0.295 | 0.91 (0.76, 1.08) | 0.267 |
| Q4 (n=13380) | 0.87 (0.73, 1.04) | 0.132 | 0.87 (0.73, 1.04) | 0.117 | 0.86 (0.72, 1.03) | 0.093 | 0.86 (0.72, 1.03) | 0.102 |
| Per SD increase | 0.96 (0.67, 1.36) | 0.808 | 0.95 (0.66, 1.35) | 0.761 | 0.95 (0.66, 1.35) | 0.764 | 0.94 (0.66, 1.34) | 0.735 |
| Retinal venular tortuosity |  |  |  |  |  |  |  |  |
| Q1 (n=13395) | Reference | - | Reference | - | Reference | - | Reference | - |
| Q2 (n=13406) | 1.18 (0.98, 1.43) | 0.085 | 1.16 (0.96, 1.41) | 0.118 | 1.19 (0.99, 1.44) | 0.068 | 1.18 (0.97, 1.42) | 0.097 |
| Q3 (n=13357) | 1.31 (1.09, 1.58) | 0.004 | 1.31 (1.09, 1.57) | 0.005 | 1.32 (1.09, 1.58) | 0.004 | 1.31 (1.09, 1.58) | 0.004 |
| Q4 (n=13361) | 1.29 (1.07, 1.55) | 0.007 | 1.19 (0.99, 1.43) | 0.061 | 1.21 (1.01, 1.45) | 0.043 | 1.19 (0.99, 1.43) | 0.064 |
| Per SD increase | 1.65 (1.19, 2.28) | 0.003 | 1.38 (1.00, 1.92) | 0.051 | 1.39 (1.01, 1.93) | 0.044 | 1.36 (0.98, 1.88) | 0.063 |
|  | HR (95% CI) | P Value | HR (95% CI) | P Value | HR (95% CI) | P Value | HR (95% CI) | P Value |
| Retinal arteriolar fractal dimension |  |  |  |  |  |  |  |  |
| Q1 (n=13832) | 1.56 (1.27, 1.92) | <0.001 | 1.38 (1.12, 1.70) | 0.003 | 1.41 (1.15, 1.74) | 0.001 | 1.35 (1.10, 1.67) | 0.004 |
| Q2 (n=13832) | 1.38 (1.12, 1.70) | 0.003 | 1.24 (1.00, 1.54) | 0.045 | 1.26 (1.02, 1.55) | 0.036 | 1.23 (1.00, 1.52) | 0.053 |
| Q3 (n=13831) | 1.19 (0.95, 1.48) | 0.129 | 1.12 (0.90, 1.39) | 0.318 | 1.14 (0.91, 1.42) | 0.258 | 1.11 (0.89, 1.39) | 0.335 |
| Q4 (n=13832) | Reference | - | Reference | - | Reference | - | Reference | - |
| Per SD decrease | 2.47 (1.73, 3.53) | <0.001 | 1.98 (1.38, 2.85) | <0.001 | 2.01 (1.40, 2.89) | <0.001 | 1.90 (1.32, 2.73) | 0.001 |
| Retinal venular fractal dimension |  |  |  |  |  |  |  |  |
| Q1 (n=13832) | 1.18 (0.98, 1.43) | 0.075 | 1.25 (1.03, 1.50) | 0.020 | 1.22 (1.02, 1.48) | 0.034 | 1.22 (1.02, 1.48) | 0.033 |
| Q2 (n=13832) | 1.14 (0.94, 1.37) | 0.184 | 1.19 (0.98, 1.44) | 0.073 | 1.17 (0.97, 1.42) | 0.102 | 1.17 (0.97, 1.42) | 0.101 |
| Q3 (n=13831) | 1.08 (0.89, 1.31) | 0.449 | 1.14 (0.94, 1.38) | 0.197 | 1.13 (0.93, 1.37) | 0.231 | 1.13 (0.93, 1.37) | 0.218 |
| Q4 (n=13832) | Reference | - | Reference | - | Reference | - | Reference | - |
| Per SD decrease | 1.40 (0.99, 1.96) | 0.054 | 1.52 (1.09, 2.13) | 0.015 | 1.47 (1.05, 2.06) | 0.025 | 1.47 (1.05, 2.07) | 0.025 |
| Number of arteriolar segments |  |  |  |  |  |  |  |  |
| Q1 (n=14159) | 1.54 (1.25, 1.90) | <0.001 | 1.36 (1.11, 1.68) | 0.004 | 1.39 (1.13, 1.71) | 0.002 | 1.33 (1.08, 1.64) | 0.007 |
| Q2 (n=14434) | 1.32 (1.07, 1.62) | 0.011 | 1.16 (0.94, 1.43) | 0.179 | 1.17 (0.94, 1.44) | 0.154 | 1.14 (0.93, 1.42) | 0.214 |
| Q3 (n=13447) | 1.17 (0.94, 1.46) | 0.156 | 1.09 (0.88, 1.36) | 0.427 | 1.11 (0.89, 1.39) | 0.340 | 1.09 (0.87, 1.36) | 0.455 |
| Q4 (n=13930) | Reference | - | Reference | - | Reference | - | Reference | - |
| Per SD decrease | 2.78 (1.89, 4.11) | <0.001 | 2.18 (1.47, 3.24) | <0.001 | 2.22 (1.49, 3.30) | <0.001 | 2.07 (1.39, 3.08) | <0.001 |
| Number of venular segments |  |  |  |  |  |  |  |  |
| Q1 (n=14099) | 1.45 (1.18, 1.77) | <0.001 | 1.41 (1.16, 1.73) | 0.001 | 1.38 (1.13, 1.69) | 0.002 | 1.38 (1.13, 1.70) | 0.002 |
| Q2 (n=14453) | 1.41 (1.15, 1.72) | 0.001 | 1.36 (1.11, 1.67) | 0.003 | 1.34 (1.10, 1.65) | 0.004 | 1.35 (1.10, 1.65) | 0.004 |
| Q3 (n=13684) | 1.23 (1.00, 1.52) | 0.053 | 1.22 (0.99, 1.51) | 0.067 | 1.20 (0.97, 1.48) | 0.093 | 1.21 (0.98, 1.50) | 0.078 |
| Q4 (n=13734) | Reference | - | Reference | - | Reference | - | Reference | - |
| Per SD decrease | 1.89 (1.32, 2.71) | 0.001 | 1.85 (1.29, 2.64) | 0.001 | 1.79 (1.25, 2.56) | 0.002 | 1.77 (1.24, 2.54) | 0.002 |

Model 1: adjusted for sex and age. Model 2: adjusted for age, sex, SBP, TC, HDL-c, TG, BMI, smoking and use of antihypertensive medication. Model 3: Model 2 plus history of stroke and history of CHD. Model 4: Model 2 plus diabetic retinopathy and RVO.

As a categorical variable, venular tortuosity in the third quartile was related to an increased risk of HF compared with the first quartile (HR, 1.31; 95% CI, 1.09-1.58; *P*=0.004) after adjusting age and sex. Even after multivariable adjustments, including retinopathy (diabetic retinopathy and RVO) and a history of stroke or CHD, these relationships were still significant. When comparing the fourth with the first quartiles, the connection between venular tortuosity and HF was statistically significant in model 1 (HR, 1.29; 95% CI, 1.07-1.55; *P*=0.007) and model 3 (HR, 1.21; 95% CI, 1.01-1.45; *P*=0.043). Similar results were also found as a continuous variable (Model 1: HR, 1.65; 95% CI, 1.19-2.28; *P*=0.003. Model 3: HR, 1.39; 95% CI, 1.01-1.93; *P*=0.044, per SD increase). We didn’t find any associations between retinal arteriolar tortuosity and the risk of HF.

For retinal vessel complexity parameters, a smaller number of vascular segments and smaller arterial fractal dimension in both arteries and veins were risk factors for HF, with a 1.36-fold greater risk in the number of arterial segments, a 1.41-fold more significant risk in the number of venous segments, and 1.38-fold greater risk in arterial fractal dimension when first quartile compared with fourth quartile in Cox proportional-hazards models adjusting for traditional risk factors. After further adjustment for retinopathy and a history of stroke or CHD, the results remained similar. When modeled as continuous variables, for 1 SD reduction in the number of arterial segments, the risk of heart failure increased by 115% (Model 2: HR, 2.18; CI, 1.47-3.24; *P*<0.001. Model 3: HR, 2.22; CI, 1.49-3.30; *P*<0.001. Model 4: HR, 2.07; CI, 1.39-3.08; *P*<0.001, per SD decrease). There was an increase in the risk of HF by 85% and 98% in the number of venous segments and arterial fractal dimensions. There was no connection between the venular fractal dimension and HF if we only adjusted for age and sex. However, after adjusting for traditional risk factors, a larger venular fractal dimension was associated with the risk of HF (HR, 1.52, 95% CI 1.09-2.13; *P*=0.015).

We performed subgroup analyses among gender, hypertension, diabetes, history of stroke, and history of CHD (STable1-8). The association of several retinal vessel measures (venous calibre, venular tortuosity, arterial/venular fractal dimension, number of arterial/venous segments) with HF was strongest in men and non-significant in women among retinal vessel measures. Veins with a wider diameter and greater tortuosity were linked to a higher risk of HF in those over 65 but not those under 65. In contrast, patients below 65 with larger venular fractal dimensions had a higher risk of HF. Lower arteriolar fractal dimension and a smaller number of vascular segments increased the risk of HF in middle-aged and elderly people. Similar results were found among participants with diabetes mellitus and a history of stroke or CHD. Venular calibre and fractal dimension were significant only in the population with diabetes mellitus or a history of stroke. Interestingly, vein diameter was significant in people without a history of CHD, whereas similar results were not found in people with a history of CHD. Except for the tortuosity index, the other six variables were linked to a higher risk of HF in the non-hypertensive cohort compared to the hypertensive population. For smokers, larger arteriolar calibre and a smaller number of vascular segments were linked to a higher risk of HF. The arterial fractal dimension was statistically significant regardless of the smoking status. Although we didn’t find an association between arterial diameter as a continuous variable and risk of HF, the further subgroup analyses indicated that the non-hypertensive population (HR, 1.64; CI, 1.11-2.44; *P*=0.014, per SD increase) and CHD-free population (HR, 2.16; CI, 1.09-4.28; *P*=0.028, per SD increase) demonstrated an association between retinal arterial calibre and HF.

### Discrimination capability of retinal vascular measures

Table 3 presented the C-Statistic for multivariate Cox regression models that predict HF in the overall sample. The C-Statistic for discrimination of HF prediction using traditional risk factors alone was 0.799 (Model A). Adding the selected retinal measures (Model B) slightly increased the discrimination capability to XXX but no statistical significance was found. Adding all retinal measures (Model C) to Model A significantly increased the performance (C statistic: Model A, 0.799; Model 0.801; *P*=0.029). Further addition of retinopathy and history of CVD made C-statistic went up to 0.812. Model F which combined all the predictors achieved a C statistic of 0.813.

**Table 3:** C-Statistic for cox regression models predicting heart failure

| Whole Model Sample | C-Statistic for Heart Failure | *P* Value |
| --- | --- | --- |
| Model A: traditional risk factors (age, sex, SBP, TC, HDL-c, TG, BMI, smoking and use of antihypertensive medication) | 0.799 | Reference |
| Model B: traditional risk factors + retinal venular calibre + retinal arteriolar fractal dimension + number of arteriolar/venular segments | 0.800 | 0.117 |
| Model C: traditional risk factors + all retinal measures | 0.801 | 0.029 |
| Model D: traditional risk factors + all retinal measures + retinopathy (diabetic retinopathy+RVO) | 0.802 | <0.001 |
| Model E: traditional risk factors + all retinal measures + history of stroke +history of CHD | 0.812 | <0.001 |
| Model F: traditional risk factors + all retinal measures + retinopathy (diabetic retinopathy+RVO) + history of stroke +history of CHD | 0.813 | <0.001 |

## Discussion

Through this study, we verified and provided additional knowledge of the association between retinal venular calibre and heart failure. Meanwhile, two new variables, fractal dimension and the number of vascular segments, were also found to be related to the risk of heart failure.

We used the mean of all venous or arterial calibre to describe retinal vessel calibre rather than CRAE and CRVE. Unlike this study’s results, an 18-year cohort study showed a higher risk of HF among participants with wider CRVE and narrower CRAE^11^. The associations were non-significant after adding SBP to the multivariate model. Different protocols used to quantify the vessel diameter may lead to different results, suggesting that all retinal vessels were dilated except the central zone. This change was more pronounced, which might better predict the risk of HF. At the same time, the calibre changes of the vessels in the central area may differ from those in the peripheral arteries. These hypotheses require further research in pathology and physiology. For arteries, the results showed that the risk of HF was reduced by 19% in the second quartile compared with the first quartile after adjusting for conventional risk variables. Still, there was no significant difference when analysing as a continuous variable. This contradicted the finding that the narrower the artery, the higher the risk of HF^11^. The contradiction may be due to different changes in the arteries in other retinal areas, which needs further research.

Besides retinal vascular diameter, we assessed several new retinal vascular characteristics in this study. To evaluate new risk factors of HF, we measured the geometric branching network of the retinal vasculature, which includes fractals, tortuosity, and several segments. Fractal dimension and the number of segments are indicators of vasculature complexity. Our results reflected that smaller fractal dimensions and the number of segments led to a higher risk of HF. The decreased fractal dimension and the number of vessel segments is a mark of vessel rarefaction and collapse^8^. Several studies showed that these changes are associated with hypoxia^22^, which would lead to microcirculation disturbance, directly affect microcirculation perfusion, and further cause the corresponding coronary artery disease, thus aggravating the deterioration of heart function and eventually leading to HF^6^. In addition, we also found that venular tortuosity was associated with the risk of HF. Elevated vessel tortuosity generally manifests as vessel wall dysfunction and blood-retinal barrier damage^23,24^. The dysfunction of retinal microvasculature, in turn, affects the blood flow regulation of the microcirculation and leads to an increased risk of local ischemia^25^, which may finally result in HF. However, the association was not significant in arteries. The reason for the discrepancy may be related to different vessel traits. Because the thinner wall of the veins is more susceptible to hemodynamic changes^25,26^.

This study found that the association with heart failure was strongest in women and non-significant in men among retinal vessel measures. Several studies have shown that females were predisposed to HF with preserved ejection fraction (HFpEF), which is closely related to microvascular function. In contrast, males are predominated affected by HF with reduced ejection fraction (HFrEF), attributed to macrovascular coronary artery disease and myocardial infarction^1,21^. In addition, pathological studies have further explained these differences. Different reproductive hormones mediate different inflammatory responses, which have different effects on regional myocardial perfusion and vasodilation^27,28^. Our findings indicated that wider venular calibre, tortuosity, and reduced fractal dimension were related to a higher risk of HF in persons over 65. Several research findings have shown age-related changes in retinal vascular parameters^29-31^. Previous studies hypothesized that this connection might be caused by the retinal vasculature’s rarefaction with aging, similar to other organ systems of the body^31-33^. Meanwhile, our research revealed that changes in retinal vascular variables were linked to incident HF among the non-hypertensive population. Previous studies revealed that people with hypertension had wider venules and reduced fractal dimensions^7^. Despite the changes in retinal vasculature in hypertensive patients, our study suggested that retinal microvasculature undergoes insidious changes before the onset of macrovascular symptoms and HF.

Our study further discussed the clinical predictive models. there appears to be an incremental value after the addition of all retinal vascular variables, retinopathy (diabetic retinopathy and RVO), and history of stroke/CHD to the traditional risk factors for HF prediction. Adding retinal measures can improve risk stratification for identifying high-risk HF patients. Nevertheless, risk prediction models still need to be optimized through in-depth research. Future studies should take into account the cost-effectiveness and acceptability and implement clinical practice to confirm the most accurate and efficient predictive models suitable for different patient subgroups.

Strengths of this study included a prospective design with extended follow-up of a large cohort, using an artificial intelligence system (RMHAS) to calculate more parameters and assess HF with validated records. Our study also has some limitations. First, we did not assess or confirm different types of heart failure and total heart failure as the outcome. Studies have shown that retinopathy and other noncardiac microvascular complications led to a higher risk of HF with HFpEF than HfrEF in diabetic patients^34-36^. It may be speculated that different types of HF had different sensitivities to retinal microvascular changes. Second, fundus images in UK Biobank were all macula centered, the disc area was not properly exposed, therefore some images were hard to calculate CRAV/CRVE, which need images taken in the disc center. Third, the present analyses were carried out in the UK Biobank, which was mostly made up of Europeans. Additional analyses in a variety of ethnic cohorts are required. In addition, our images and methods need to be revised to reflect deep vessel parameters. Recent research has revealed that the retinal superficial and deep blood vessels are all regulated differently at the molecular and cellular levels^37^. Thus, further study can pay attention to superficial and deep vessels to explore the pathophysiological mechanism.

In summary, our study showed that retinal venular calibre, arteriolar fractal dimension, and the number of vessel segments were associated with incident heart failure events. These results point to the possibility and usefulness of using early retinal vasculature changes to identify people at higher risk of incident heart failure.

## Data Availability

Data from the UK Biobank are available to researchers by application via the UK-Biobank online Access Management System.

